# Novel approach reveals lipid metabolite reduction in nails of breast cancer patients as potential biomarker

**DOI:** 10.1101/2020.04.14.20064675

**Authors:** Manmohan Mitruka, Charusheela R. Gore, Ajay Kumar, Sachin C Sarode, Nilesh Kumar Sharma

**Affiliations:** Cancer and Translational Research Lab, Dr. D.Y. Patil Biotechnology & Bioinformatics Institute, Dr. D.Y. Patil Vidyapeeth, Pune, Maharashtra, India, 411033; Department of Pathology, Dr. D. Y. Patil Medical College, Hospital and Research Centre, Pimpri, Pune, Maharashtra, India; Department of Oral Pathology and Microbiology, Dr. D.Y. Patil Dental College and Hospital, Dr. D.Y. Patil Vidyapeeth, Pimpri, Pune, India

**Keywords:** Neoplasms, Metabolic reprogramming, Microenvironment, Diet, Free aromatic amino acids, Biomarker

## Abstract

**BACKGROUND:** Molecular adaptations in intracellular and extracellular microenvironment of breast cancer cells promote pro-tumor metabolic reprogramming. Hence, metabolic reprogramming is seen as a crucial factor in various tumor hallmarks including drug resistance, invasiveness and metastasis. Among well-known metabolic features of breast carcinoma including Warburg effects, altered amino acid metabolism, lipid remodeling is considered as key factors in achieving pro-tumor microenvironment. Therefore, a better understanding on molecular aspects of lipid remodeling is highly appreciated that may contribute towards future therapeutics and diagnostics purpose including the need of potential biomarkers. The identification and validation of lipid biomarkers are reported in the literature, but evidence on lipid metabolites as biomarkers in nails of breast cancer patients is completely unexplored.

**METHODS:** This study reported a novel and specifically designed vertical tube gel electrophoresis (VTGE) system to assist in the purification of metabolites in the range of (∼100-1000 Da) from nail samples. Fingernail clippings of breast cancer patients (N=10), and healthy subjects (N-12) were used for extraction and purification of metabolites. The VTGE system uses 15% polyacrylamide under non-denaturing and non-reducing conditions that makes eluted metabolites directly compatible with LC-HRMS and other analytical techniques. The characterization of lipid metabolites in nail lysates was done by positive ESI mode of Agilent LC-HRMS platform.

**RESULTS:** Data suggest a novel observation that healthy and breast cancer patients show distinct accumulation of lipid metabolites specifically choline-based lipids. This is a first report that suggests that levels of choline, phosphorylcholine and lyso-PC are highly reduced and undetectable in nails of breast cancer patients over healthy subject. Furthermore, the potential use of reduced level of choline, phosphorylcholine and lyso-PC in nails of breast cancer patients is in line with current notion that these lipids are diverted to meet the pro-tumor activities in the tumor microenvironment.

**CONCLUSION:** Data strongly provide a proof of concept for the potential use of lipid metabolites including choline, phosphorylcholine and lyso-PC as a set of biomarkers in nails of breast cancer patients. However, the authors propose that validity of these lipid biomarkers may be extended to large population size of breast cancer patients for future applications in early detection, grading, staging, predicting prognosis and therapeutic targeting of breast carcinoma.

## INTRODUCTION

With increasing burden of breast cancer incidences at global level, around 1.5 million of new cases of breast cancer is projected each year (1-2). Therefore, comprehensive and novel approaches are warranted to meet the requirements of early diagnosis, monitoring of drug response and new modalities of treatment options (3-6). Therefore, new area of genomics, proteomics, transcriptomics, epigenomic and metabolomics is set to make advancement in exploring novel classes of biomarkers that can assist in the early detection and drug response monitoring avenues in a global context and Indian setting (3-10).

The importance of metabolic heterogeneity is seen as one of key factors that contribute towards biomarkers development in breast cancer patients (5-11). Metabolic heterogeneity encompasses various metabolic adaptations including lipid remodeling by breast cancer cells to suffice the metabolic requirements in the tumor microenvironment (9-17). Therefore, appreciable attempts were made to determine lipid profiles mostly in blood, as it is the viable option to detect early and late stage physiological and pathological changes in breast cancer patients (10-17). Among key lipid metabolites, levels of choline derived lipids such as phosphatidylcholine (PC), phosphorylcholine and lyso-PC are suggested to decrease in breast cancer and other cancer types and these metabolites are seen as potential biomarkers (13-14,18-21). In the current approach of lipid metabolomics, LC-HRMS, GC-MS and NMR spectroscopy were widely employed on whole metabolite extracts from blood and other biological fluid samples (10-21).

Besides the use of traditional biological sources, an alternative use of nails as a source of biomarker discovery in breast cancer patients and other pathological condition is highly limited. However, limited attempts are available that investigated the nail metabolite profiling in an environmental exposure cases and disease conditions (22-25). Till date there are no suitable methodology that targets to profile nail metabolites of breast cancer and other tumor types. Furthermore, the importance of nail over other biological fluids can be justified in terms of issues related to contamination from microbes and other environmental factors. Moreover, other biological fluid/material shows high degree of diurnal variation due to human physiology and dietary intakes. At the same time, nails may show the stable concentration of metabolites over a long period of accumulation of metabolites including lipids and thus lacks the diurnal variation.

In this paper, we report a novel and specifically designed vertical tube gel electrophoresis (VTGE) based lipid metabolite identifications in nail lysates. Further, data reported that reduced/undetectable levels of choline, phosphorylcholine and lyso-PC in nail lysate of breast cancer patients may serve as a potential set of metabolite biomarker.

## Methods

### Study Population

Breast cancer patients (n=10) and healthy women subjects (n=12) were recruited from Dr. D. Y. Patil Medical College and Research Center, Pune, India. Institutional Ethics Committee approval was obtained before commencement of the study. The IEC is named as Ethics Committee of Dr. D. Y. Patil Vidyapeeth, Pune and this is affiliated to Dr. D. Y. Patil Vidyapeeth, Pune, India with a registration no Re-Reg.No.ECR/361/Inst/MH/2013/RR-16. All the participating study population was apprised about objectives of the study and informed consent was obtained prior to investigations.

Metabolite profiling data were analyzed between breast cancer patients and healthy subjects, without grouping into molecular sub-types. The primary goals were to develop a protocol for the preparation of the nail metabolite lysate, followed by purification process of nail metabolites and then finally characterizing with the help of LC-HRMS. At this stage of study, we focused on the novel methods and processes that helped to collect metabolite profiles in the nail lysates of breast cancer and healthy subjects.

### Preparation of nail lysates

Fingernail clippings were collected in a microfuge tube and cleaned to remove debris and environmental contaminations by using a mild detergent and 70% ethyl alcohol. Further, fingernail clippings were dried, weighed and coded properly for healthy and breast cancer patients. Next, equal amount of fingernail clippings (20 mg) was added to 800 µl of extraction buffer (Tris-HCl (20mM, pH-8.5), 2.6M Thiourea, 5M Urea) and 800 µl Beta-Mercaptoethanol) for lysis of fingernail clippings. Sample mixture was incubated for 24 hr at 50LJC under dark condition and then centrifuged at 15,000 X RPM for 30 min. The supernatant was collected in a fresh microfuge tube and filtered by using 0.45 micron syringe filter membrane. The above prepared nail lysates of healthy and breast cancer patients were diluted three times, stored and labeled properly for purification by using Vertical Tube Gel Electrophoresis (VTGE) system.

### Purification of metabolites by using Vertical Tube Gel Electrophoresis system

In order to purify metabolites of nail materials, a novel and specifically designed VTGE system was used. It was standardized to use 15% polyacrylamide gel matrix to remove major components as proteins, polysaccharides, large lipid molecules and other various debris/contaminants. At the same time, this novel VTGE system allowed the purification of metabolites in the range of ∼100 Da to 1000 Da (26-29).

Above prepared sterile and filtered nail lysate (750 µl) was mixed with (250 µl) the loading buffer (4X Glycerol and Tris pH-6.8) and electrophoresed on VTGE casted with 15% acrylamide gel (acrylamide: bisacrylamide, 30:1) as a matrix. The fractionated elute was collected in electrophoresis running buffer that contains water and glycine, and excludes traditional SDS, and other reducing agents. A notable distinctiveness of VTGE system is elaborated on the use running buffer (3 ml) and elution buffer (3 ml), both are identical as (5X Water-Glycine (192 mM), pH: 8.3). A flow diagram of VTGE method is presented in Figure 1A and 1B that shows the assembly and design of VTGE system. Nail lysate metabolites from healthy and breast cancer patients were eluted in the same running buffer, which is referred as “Elution buffer”. It was placed in the lower electrophoretic tube that has anode wire. Voltage and current ratio from power supply was maintained to generate 1500-2500 mW of power to achieve the electrophoresis of biological samples. The total run time was allowed for 2 hr. which was then collected in a fresh microfuge tube for direct LC-HRMS characterization. Interestingly, pH of eluted metabolite buffer was measured for healthy and breast cancer patients that ranged between 3.0-3.5 acidic pH. An acidic pH (2.5-4.0) buffer containing metabolites are known to increase the ionization efficiency during LC-HRMS analysis (26,27). At the end of the run, inner tube containing polyacrylamide gel was removed and placed for coomassie brilliant blue dye staining to ensure that protein components of nail lysates were trapped in the polyacrylamide matrix. Eluted nail metabolites from healthy and breast cancer patients were stored at -20°C for further direct identification by LC-HRMS technique.

**Figure 1.**
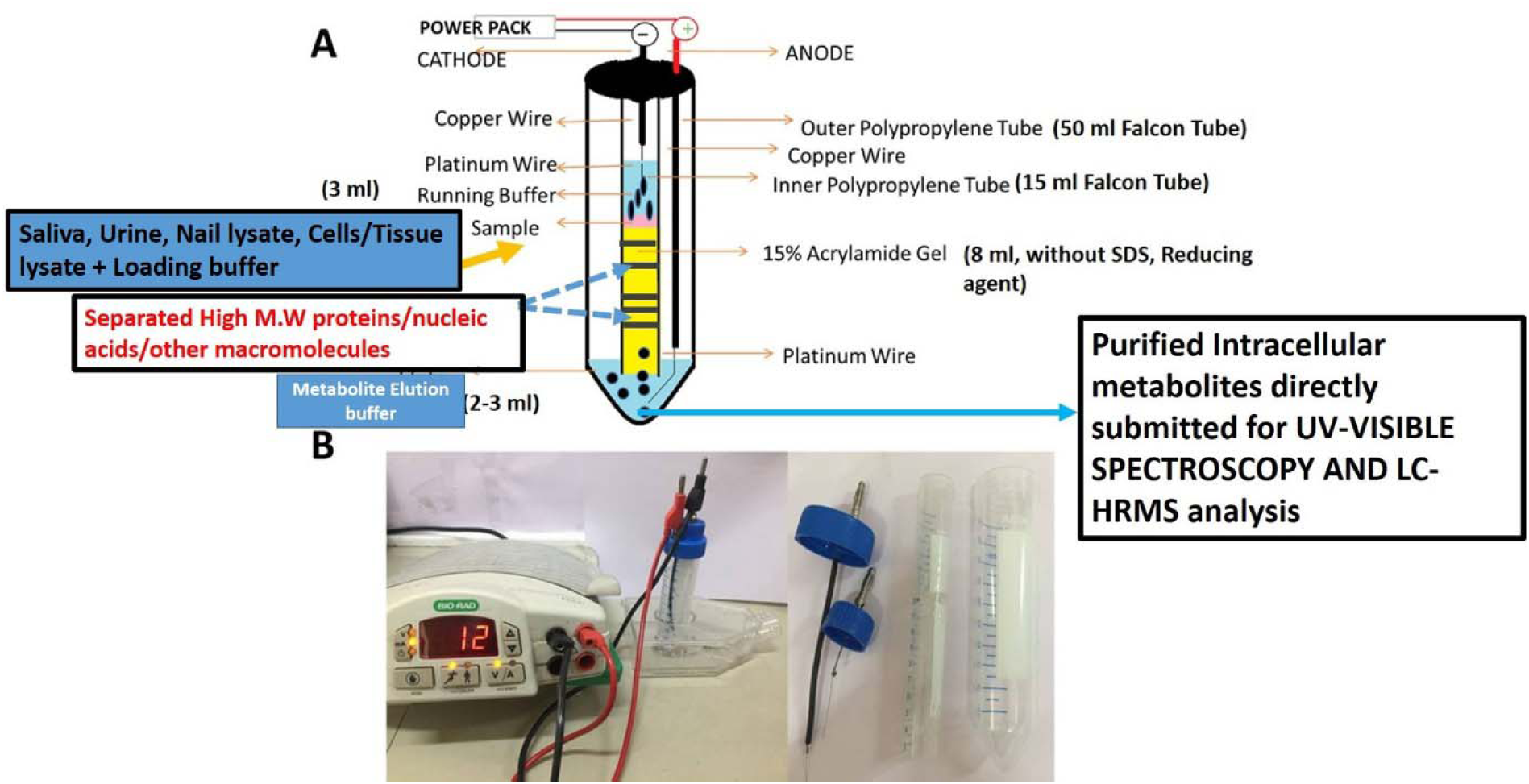
A flow diagram of novel and specifically designed vertical tube gel electrophoresis (VTGE) system for intracellular nail metabolite purification. Here, (1A) depicts the design, assembly and key features including nature of matrix, non-reducing and non-denaturing buffers. (1B) shows the working diagram that allow the nail metabolite purification from healthy subjects and breast cancer patients.

### Identification of potential nail metabolites by LC-HRMS

The purified nail metabolites from VTGE system was submitted to LC-HRMS analysis. For liquid chromatography (LC) component, RPC18 column was used as Zorbax, 2.1 X 50 mm, micron meters. Further, a flow rate of 0.2 ml/min and a gradient was formed by mixing mobile phase A (water containing 5mM ammonium acetate) and B (0.2% formic acid). For running the sample, an injection volume was of 25 µl and a flow rate of solvent was maintained at 0.3 ml per minute. The HPLC column effluent was allowed to move onto an Electrospray Ionization Triple Quadrupole Mass Spectrometer (Agilent Technologies). For mass identification of nail metabolites, samples were run in positive electrospray ionization (ESI) M-H mode. During LC-HRMS analysis, mass spectrometer component was used as MS Q-TOF Quadrupole time-of-flight mass spectrometry (Q-TOF-MS) (Agilent Technologies, 6500 Series Q-TOF LC/MS System) with dual AJS electrospray ionization (ESI) mode. The acquisition mode of MS1 was recorded with a minimum value of m/z at 60 and maximum value of m/z at 1700 (27-29).

## STATISTICAL ANALYSIS

Data was presented as the mean ± SD. The statistical significance between the healthy subjects and breast cancer patients were assessed with the help of students unpaired t-test. Data calculation and statistical tests were performed by SPSS version 15.0 (SPSS) software package.

## RESULTS AND DISCUSSION

The importance of metabolite adaptations and profiling of metabolite biomarkers in fine-needle aspiration biopsies, surgical biopsies, serum, urine and saliva was shown in various tumor types such as breast, liver, thyroid and colorectal cancer (9-17). Among various classes of metabolites, choline and choline-related lipids are explored as potential biomarkers in tissue and plasma of breast cancer as well as other cancer types (9-17). Besides these existing preclinical and clinical evidences, attempt to identify choline and choline-related lipid metabolite in nails of breast cancer patients is not reported by a single study. Furthermore, a suitable and efficient methodology to extract, purify and identify lipid metabolite biomarkers a nail lysate of cancer patients is completely lacking in the field of biomarker discovery

To achieve new and additional knowledge on metabolomic biomarkers in breast cancer, we developed a novel and specifically designed VTGE system that involved methods to purify nail lysate metabolites. The lysates are directly compatible with LC-HRMS techniques without any further need of complicated extractions, modifications and labeling protocols to identify and estimate metabolite biomarkers. In this paper, the reported VTGE method is simply the out-of-box innovative applications with respect to classical Laemmli (1970)^26^ system that is primarily used for the separation and purification of large macromolecules such as proteins and nucleic acids. In this novel method, the authors have designed a VTGE system with the help of laboratory plastic ware that purifies nail metabolites (∼100 Da-1000 Da) by direct elution in the lower running buffer.

Limited reports support the avenues to explore nail metabolites, including ethyl glucuronide in the keratinous matrices of nail materials of patients (22-25). In addition to choline and choline-related lipid metabolites, nail lysates of human subjects including healthy and cancer patients show abundance of other metabolites such as fatty acids, amino acids, vitamin, drug derived metabolites (24-29). Therefore, we have recently proposed that nail serve as a biological reservoir that frequently gets clipped off. Interestingly, molecular weight of these identified novel methodology exactly within the expected metabolite purification range of ∼100-1000 Da. However, there is not a single paper that addresses the relevance of choline and choline-related lipid metabolites as potential biomarkers in nail materials of breast cancer patients.

### Level of choline and phosphorylcholine in nails

There are clear understanding on the heightened choline metabolism in cancer cells and tissues that support the progression, invasion and metastasis of cancer cells (9-17). In other way, cancer cells are known to divert the dietary source including choline to support the metabolic needs of breast carcinoma and other cancer cell types (11-16). There are preclinical and clinical findings that support the high abundance of cellular choline and choline based metabolism in cancer cells. On the other hand, membrane bound PC, lyso-PC and phosphorylcholine are reduced in the extracellular fluids including plasma (12-21). In essence, distinct lipid metabolite pools at intracellular and extracellular levels are suggested to be a concerted efforts between cancer cells and associated tumor microenvironment that support abundance of various lipid metabolizing enzymes that modulate the metabolite profile as pro-tumor (11-15). Here, we showed the clear and novel evidence on the appreciable reduction of choline and phosphorylcholine in the nail lysate of breast cancer patients compared to healthy subjects (Table 1 and Table 2). A representative positive EIC of choline (+ESI EIC 86.096, 104.1070) and phosphorylcholine (+ESI EIC 185.0811) is illustrated in Figure 2 and Figure 3, respectively. The relative arbitrary abundance by LC-HRMS of choline and phosphorylcholine in nail lysate of healthy subject is found to be at 21303 ± 3810 and 45275 ± 9652, respectively. Conversely, data suggest the undetectable level of choline and phosphorylcholine in the nail lysate of breast cancer patients. The highly efficient mass ionization and quantitative analysis of choline and phosphorylcholine is obtained in purified metabolites of nails and success of these results are attributed to the novel purification methods assisted by VTGE system.

**Table 1.** List of nail lipid metabolites and their abundance in nail lysates

| Sr. No | Name of Free aromatic amino acids | Healthy subjects* | Breast cancer patients |
| --- | --- | --- | --- |
| 1. | Choline | 21303 ± 3810 | Undetectable |
| 2. | Phosphorylcholine | 45275 ± 9652 | Undetectable |
| 3. | LysoPC(16:1(9Z)/0:0) | 45933 ± 7704 | Undetectable |

\* Data represent mean ± S.D (N=10 Breast cancer patients and N=12 for healthy subjects)

**Table 2.** List of free aromatic amino acids and their abundance in nail lysates

| Sr. No | Name of nail metabolites FAAAs and metabolites | Formula | +ESI EIC | m/z | Mass | Polarity |
| --- | --- | --- | --- | --- | --- | --- |
| 1 | Choline | C5 H14 N O | 86.096<br>104.1070 | 86.0967 | 104.1078 | Positive |
| 2 | Phosphorylcholine | C5 H15 N O4 P | 185.0811 | 185.0812 | 184.0741 | Positive |
| 3 | LysoPC(16:1(9Z)/0:0) | C24 H48 N O7 P | 516.3061 | 493.3168 | 493.3168 | Positive |

**Figure. 2.**
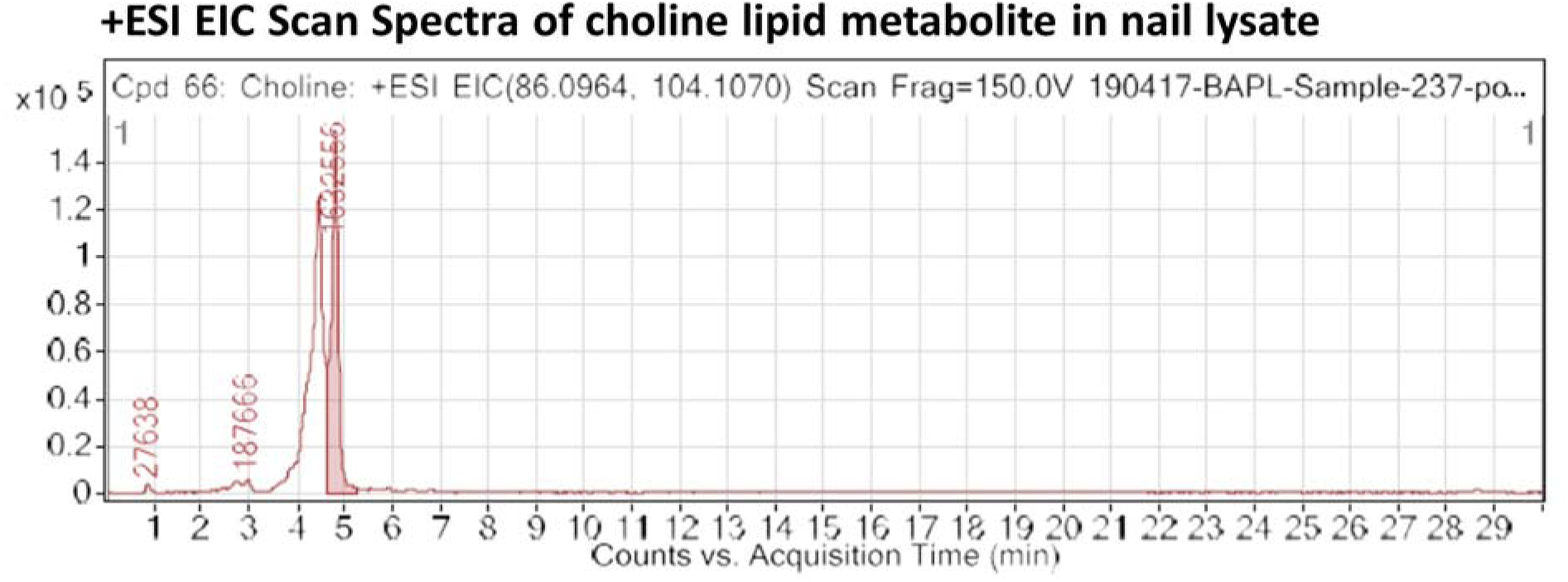
A lipid metabolite choline is abundant in nail lysate of human subject through metabolite profiling analysis by LC-HRMS. An extracted ion chromatogram (EIC) of choline, a lipid metabolite shows a +ESI EIC with a characteristic mass ion 86.0964 in nail lysates. This EIC spectra is obtained from LC-HRMS analysis of nail lysates purified by a novel and specifically designed VTGE metabolite purification system.

**Figure. 3.**
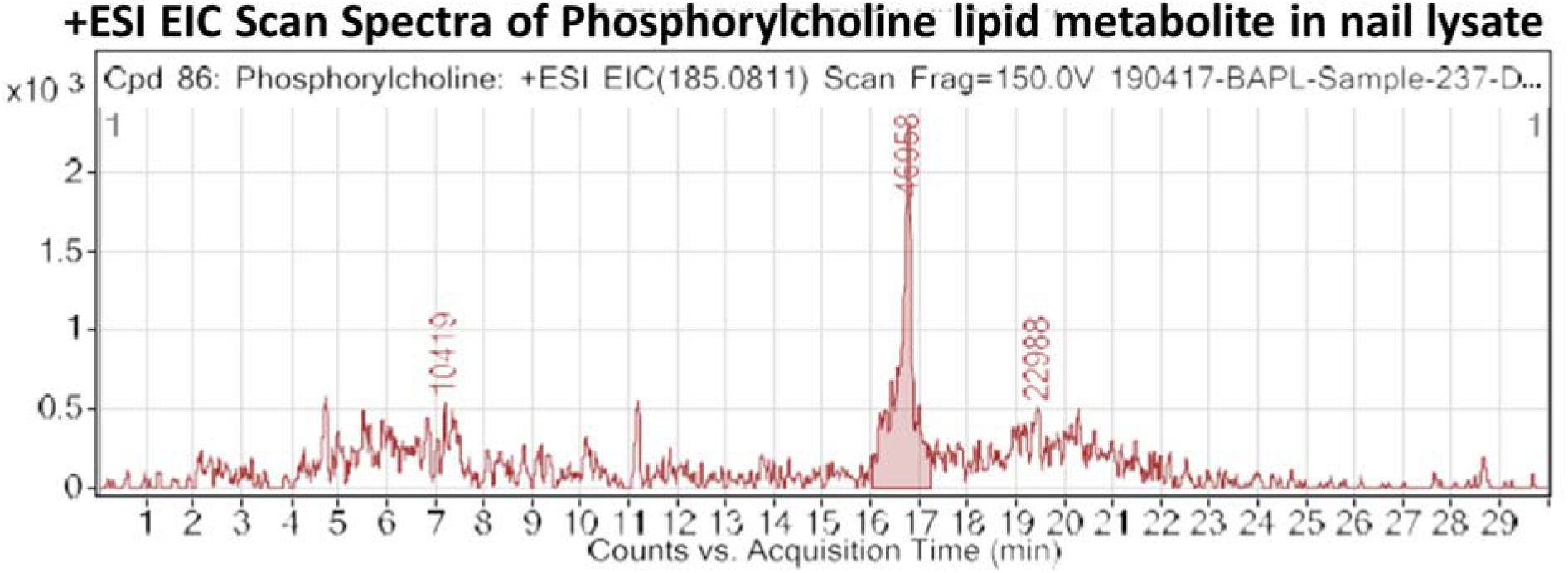
Metabolite profiling analysis of nail lysates by LC-HRMS show the abundance of phosphorylcholine lipid metabolite. An extracted ion chromatogram (EIC) of phosphorylcholine, a lipid metabolite shows a +ESI EIC with a characteristic mass ion 185.0811 in nail lysates. This EIC spectra is obtained from LC-HRMS analysis of nail lysates purified by a novel and specifically designed VTGE metabolite purification system.

Malignancy of cancer cells are known to emanate from genetic alterations that set the stage for pro-tumor metabolism of choline phospholipids leading to mitgenic signals (3-7). In other ways, intracellular accumulation of choline phospholipids, phosphorylation of choline is a key feature of breast cancer tissue and suggested as potential diagnostic marker (3-7). An interesting LC-MS study in the liquid biopsies of breast cancer patients substantiate that cells accumulate high level of choline and choline-related biomarkers such as phosphoryl choline (16). A cell-based study provides evidence on the high accumulation of phosphorylcholine in breast cancer cells compared to normal epithelial cells. Indeed, high level of phosphorylcholine in breast cancer cells are linked to enhanced expression of choline kinase enzyme that converts choline into phosphorylcholine (13,18). A study by proton magnetic resonance spectroscopy (1)H MRS) suggests that a higher metabolic conversion of membrane phosphatidylcholine to choline and phosphocholine in breast cancer cells (13,15). Such observations are associated with the overexpression of choline kinase and phospholipase C in breast cancer cells that enable them to achieve pro-tumor lipid remodeling. Such study strengthen the possibility of reduced accumulation of choline and choline-related metabolites in the plasma and other biological fluids (9-16). In the present finding, we clearly show that nails of breast cancer patients show the reduced or almost undetectable level of choline and phosphorylcholine, which is in line with previous findings reported on lipid biomarkers in plasma and tissue samples. Existing views on altered choline and choline-related lipid metabolism as a hallmark of breast cancer is strengthened by an crucial evidence that support that breast cancer patients have undetected levels of choline and phosphorylcholine in nails compared to healthy subjects.

### Undetectable level of lyso-PC in nails of breast cancer patients

Cancer cells are equipped with metabolic machinery to remodel the lipid profiles in the intracellular and extracellular compartments that favors the pro-tumor microenvironment (13-21). There are significant findings that indicate the healthy human plasma has abundance of lysophophatidylcholine (LysoPC) that is required for the various physiological needs (13-21). Conversely, patients with malignant cancer diseases are suggested to show the low level of LysoPC in plasma levels and proposed as potential metabolite biomarkers (18-21). In this paper, data obtained by LC-HRMS analysis of purified nail metabolite provide a clear difference in the level of lyso-PC between healthy subjects and breast cancer patients. In case of nail lysate from healthy subject, level of lyso-PC is biologically relevant an arbitrary unit at 45933 ± 7704, compared to undetectable level in breast cancer patients (Table 1 and Figure 4). The positive EIC of detected lyso-PC in nail lysate shows a characteristic mass ion of 516.3061 and this matches with existing databases and previous studies (13-21).

**Figure. 4.**
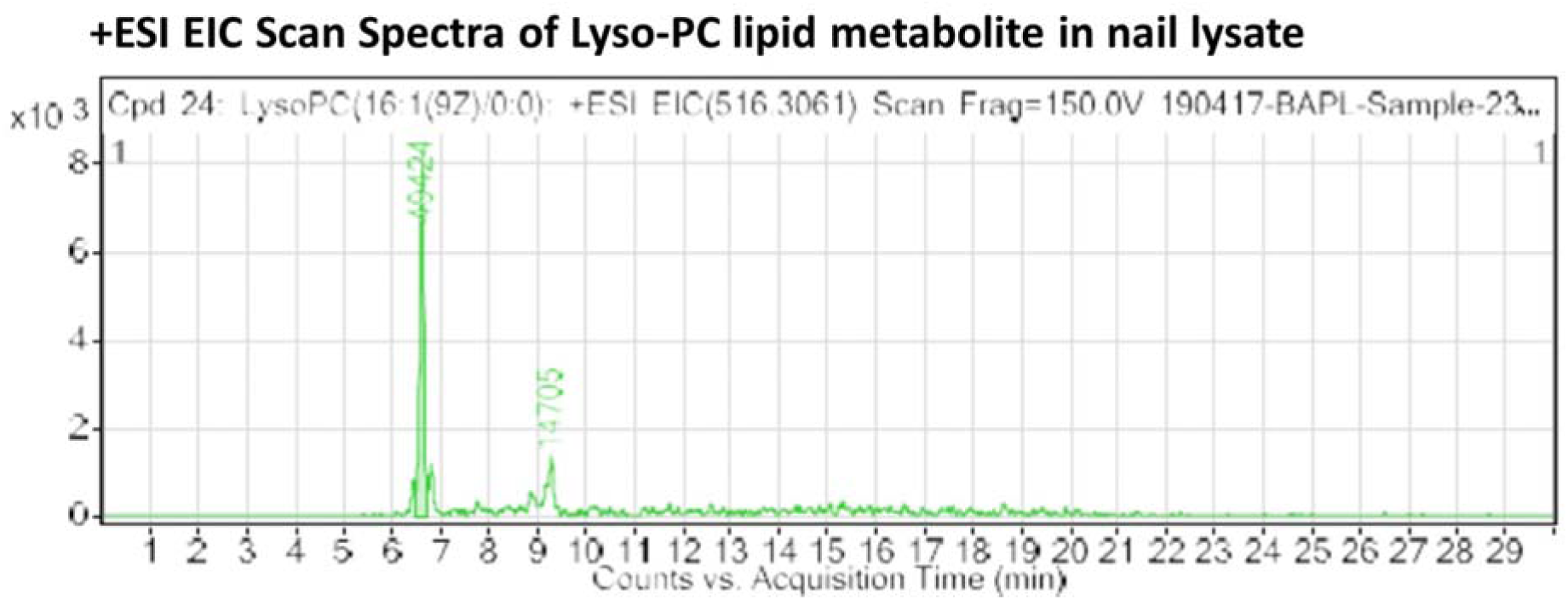
LC-HRMS analysis of nail lysates of selected human subjects indicates abundance of Lyso-PC lipid metabolite. An extracted ion chromatogram (EIC) of Lyso-PC, a lipid metabolite shows a +ESI EIC with a characteristic mass ion 516.3061 in nail lysates. This EIC spectra is obtained from LC-HRMS analysis of nail lysates purified by a novel and specifically designed VTGE metabolite purification system.

Sufficient clinical studies suggest the decrease of choline-related lipid degradation product lysoc-PC in the plasma of cancer patients including breast, colorectal cancer (13-21). Such study supports the existing view on the capability of tumor to achieve lipid remodeling for desired progression and invasiveness. Furthermore, a prospective metabolomics study reveals that high level of lyso-PC is associated with the reduction in the occurrence of common cancers (21). In the context of clinical observations that suggest the reduction in level of lyso-PC in the plasma of cancer patients. A study suggests that overexpression of lysophosphatidylcholine acyltransferase (LPCAT1) is associated with the abnormal lipid remodeling that support the growth requirements of cancer cells mediated by the oncogenic growth factor receptors (18-21). Hence, existing views and evidences on reduced level of lyso-choline in plasma of breast cancer patients raise strong possibilities that nails may accumulate highly reduced lyso-PC. Interestingly, this paper provide a first proof of concept evidence on the use of nails as a source biological material that will serve in future for metabolite biomarkers in breast cancer patients.

A prominent importance of this paper is the use of novel and specifically designed methodology that assisted to extract metabolites from nail lysates of human subjects and precisely purified by VTGE system. Importantly, purified metabolites of nail lysate were obtained in an eluting buffer that shows high efficiency and reproducibility during LC-HRMS identifications. In literature, reported approaches for plasma and other biological fluids involved extraction method to prepare the extracts that potentially contains numerous biological components including proteins, peptides, and large macromolecules that creates potential difficulties in analysis by LC-HRMS and NMR spectroscopy. Therefore, this paper is first report that explores the potential of reduced level of choline and choline-related metabolites such as phosphorylcholine and lyso-PC in nails of breast cancer patients over healthy control.

In the present study, breast cancer patients were not characterized into grades, stages and molecular profile. The lipid metabolite values might differ according to variations in these parameters and hence future studies are recommended in this direction. Second limitation of the study is less sample size due to availability of the limited resources. However, sample size is enough to establish proof of concept for future studies. For projecting the present methodology as diagnostic, prognostic or therapeutic marker, a study on larger sample size is warrented.

## CONCLUSION

Metabolic reprogramming is a common feature of cancer progression and metastasis. Besides the Warburg effect, tumour cells also undergo lipid remodeling mostly characterized by changes in the lipid profiles based on choline and choline-related metabolites. However, current findings bring an additional and novel proof of concept on the reduced level of choline and choline-related metabolites such as phosphorylcholine and lyso-PC may serve as potential biomarkers for breast cancer. However, the described novel methodology has applications in general for other cancer types to reveal the novel metabolite biomarkers due to suitability and dedicated features for metabolite biomarkers study. It is first of its kind novel proof of concept study on breast cancer metabolomic in nail lysate that showed the reduced level of choline, phosphorylcholine and lyso-PC as a potential set of metabolite biomarkers. Furthermore, the described methods and process warranted future studies to establish metabolite biomarker in other biological fluids/tissues in various cancer models.

## Data Availability

Yes Data will be made available against the request.

## ACKNOWLEDGEMENTS

The authors acknowledge financial support from DST-SERB, Government of India, New Delhi, India (SERB/LS-1028/2013) and Dr. D.Y. Patil Vidyapeeth, Pune, India (DPU/05/01/2016).

---

**Figure 5.**
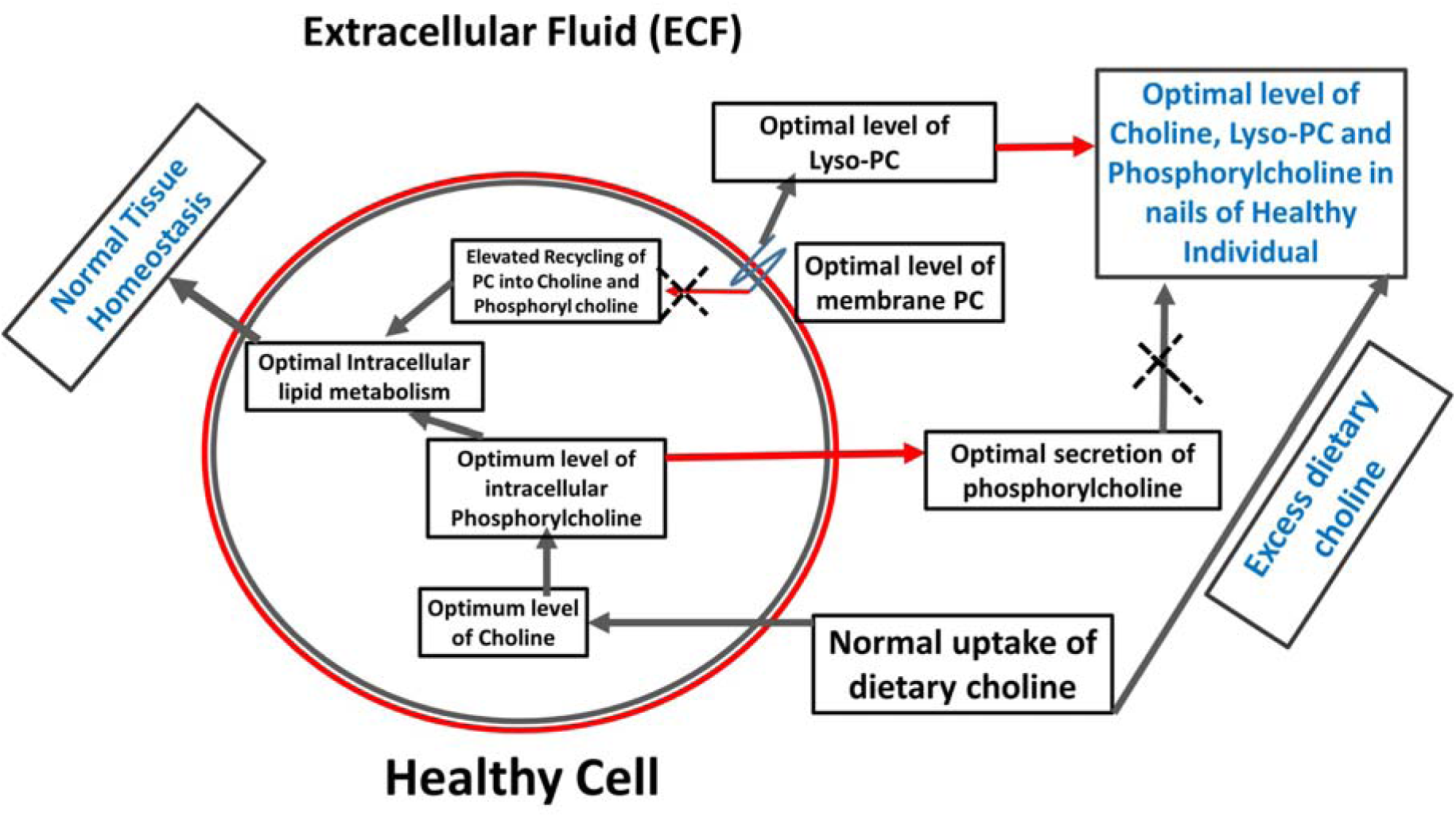
A proposed model that elucidate the abundance of choline and choline related lipids in the nails of healthy subject. This figure proposes a model to explore the choline and choline related lipids as abundant metabolites in nails of healthy subjects. Here, choline, phosphorylcholine and lyso-choline are accumulated in the nails of healthy subject.

**Figure 6.**
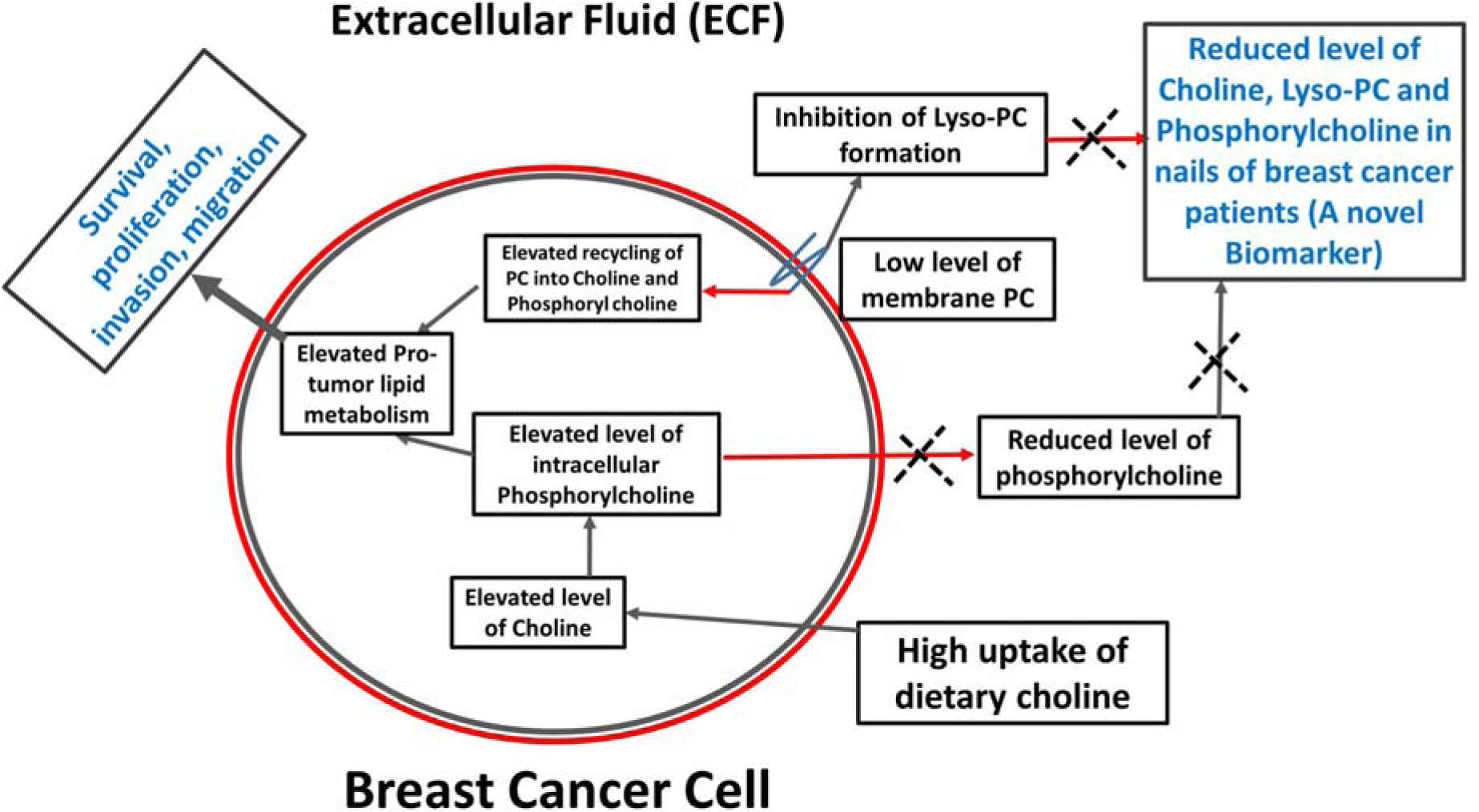
A proposed model that show choline and choline-related lipids are highly reduced in nails of breast carcinoma patients. Due to altered lipid remodeling in breast cancer patients, the level of choline and choline-related lipids including phosphorylcholine and lyso-PC are undetectable in the nails of breast cancer patients compared to the healthy subjects. These lipid metabolites are proposed as a potential metabolite biomarkers for the early detection of breast cancer patients.

